# Opportunities to Address Specialty Care Deserts and the Digital Divide: VA’s Virtual Hub-and-Spoke Cardiology Clinic

**DOI:** 10.1101/2023.10.17.23297184

**Authors:** Rebecca Tisdale, Colin Purmal, Neil Kalwani, Alexander Sandhu, Paul Heidenreich, Donna Zulman, Tanvir Hussain

## Abstract

**Background:** Access to specialty care, including cardiology, in the Veterans Health Administration (VHA) varies widely across geographic regions. VHA’s clinical resource hub (CRH) model of care offers mostly-virtual specialty care to individuals in low access regions and has recently been implemented in cardiology. How implementation of this predominantly virtual cardiology program affects the reach of cardiology specialty care in VHA is not known. This study describes the association between patient characteristics and use of CRH cardiology care in VHA’s Sierra Pacific region (Northern California, Nevada, and the Pacific Islands).

**Methods:** We compared patients who used CRH cardiology services between 7/15/2021 and 3/31/2023 to non-CRH Sierra Pacific cardiology patients, then used multivariate logistic regression to estimate the association between patient-level factors and odds of being a CRH user.

**Results:** There were 804 CRH users over the study period with 1,961 CRH encounters, and 19,583 non-CRH users with 83,489 encounters. Among CRH users, 8% were women and 41% were ≥75 years, compared to 5% and 49% respectively among non-CRH users. Similar proportions in both groups were rural (26% for both CRH and non-CRH), highly-disabled (48% CRH, 47% non-CRH), and low-income (21% CRH, 20% non-CRH). In multivariate logistic models, adjusted odds of using CRH were higher for women (adjusted odds ratio [AOR] 1.70 [95% CI 1.46-1.98]) and lower for older Veterans (AOR 0.33 for ≥75 [95% CI 0.23-0.48]). Highly rural Veterans also had higher adjusted odds of using CRH (AOR 1.88 [95% CI 1.30-2.69]).

**Conclusions:** The Sierra Pacific CRH cardiology program served a disproportionately high number of women and highly rural Veterans and similar proportions of highly-disabled and low-income Veterans as conventional VA care in its first two years of operation. This predominately-virtual model of cardiology care may be an effective strategy for overcoming access barriers for certain individuals, though targeted efforts may be required to reach older Veterans.

## Introduction

Access to specialty care, including cardiology, in the Veterans Health Administration (VHA) varies widely across geographic regions.^1–3^ Given the high prevalence of cardiovascular disease and associated morbidity and mortality among Veterans,^4^ maintaining access to cardiology care is essential. As there are unique and disproportionately high risks of dual use of VHA and community care for Veterans with cardiovascular disease,^5^ maintaining access to VHA-based cardiology care is a particular priority.

Virtual care (new or follow-up patient visits, delivered by phone or video) expanded significantly during the COVID-19 pandemic in cardiology^6^ as across the VHA^7^ and other healthcare systems. In this post-public health emergency phase of the pandemic, patient familiarity with these modalities of care provides opportunities for new ways of using virtual care, including to improve access to specialty care in non-emergency settings.

VHA’s clinical resource hub (CRH) model of care offers mostly-virtual care to individuals in low access regions. The CRH cardiology program was first implemented in July 2021 in VA’s Sierra Pacific region, which serves Northern California, Nevada, and the Pacific Islands (see Box).

#### Box 1. VHA’s Clinical Resource Hub (CRH) program

The Palo Alto VHA site serves as the ***hub*** for specialty care, including cardiology, for VHA’s CRH program in the Sierra Pacific region. ***Spoke*** site-specific contracts known as Telehealth Service Agreements detail the relationship between the Palo Alto-based CRH clinical team and other individual sites; the spoke sites implementing CRH cardiology in 2021-2023 in this region were VA’s Sierra Nevada (Reno), Southern Nevada (Las Vegas), and Northern California (Sacramento) sites. The services available via CRH at each spoke site (e.g., which subspecialty clinics, such as heart failure or women’s health cardiology) depend on the site’s needs and service gaps, and the program employs physicians, nurses, pharmacists, and administrative staff.

While the initial implementation and usage of the CRH model has been described in primary and mental health^8,9^ and for certain specialties^10,11^, expansion of the program for cardiology specialty care has yet to be characterized. In this study, we will analyze the initial implementation of the VISN 21 CRH cardiology program, describing program growth, sociodemographic characteristics of users, and different modalities of care across the program.

## Methods

This analysis was conducted as a clinical operations quality improvement project through the VISN 21 Clinical Resource Hub leadership team and was therefore exempt from IRB approval.

Using VA’s Corporate Data Warehouse, we constructed a cohort of all patients with at least one evaluation & management (E&M) encounter in any CRH or conventional VHA cardiology clinic in VA’s Sierra Pacific region (encompassing Northern California, Nevada, and the Pacific Islands) between 7/15/2021, when the first clinical resource hub site was first implemented, and 3/31/2023.

### Patient Characteristics

We included the following patient-level sociodemographic data: age, sex, race/ethnicity (American Indian/Alaska Native, Asian, Black/African American, Native Hawaiian/Pacific Islander, White, or unknown), rurality (highly rural, rural, or urban), and home site for receiving VHA care. We also included VHA enrollment priority as a proxy for social need, based on VHA’s enrollment priority classification system;^12^ this program categorizes VHA patients according to military service-related disability and income and influences whether patients pay co-pays and what services they can access within VHA. As in prior published literature,^6,7^ we condensed these enrollment priority categories to four: high disability, corresponding to enrollment priority groups 1 and 4; low-moderate disability, including priority groups 2, 3, and 6; low-income, including priority group 5; and no disability nor low-income status, wherein patients pay co-pays for VHA care, including priority groups 7-8. Due to the hierarchical nature of these groups, Veterans assigned to high- or low-moderate disability groups may also be low-income.

We captured cardiovascular diagnoses based on primary diagnoses at cardiology visits, grouping these into several categories representing the most commonly-coded primary diagnoses: heart failure, ischemic heart disease, valvular heart disease, and atrial fibrillation/flutter. International Classification of Disease (ICD)-10 codes are shown in Table S1.

### Encounter Characteristics

We captured the primary diagnosis group assigned to a given encounter as described above, as well as the encounter hub and spoke sites. We also collected data on encounter modality: phone, video (either direct to the patient’s home via VHA’s video platform, or from the cardiology team to a local clinic), or in person.

### Statistical Analysis

We constructed a logistic regression model with primary outcome of adjusted odds of being a CRH user, with all the covariates outlined above and with clustering by the patient’s assigned primary care site. We then constructed a separate logistic regression model of adjusted odds of being a video care user.

## Results

### Patients

There were 804 CRH users over the study period with a total of 4,315 ambulatory cardiology encounters, 1,961 of which were CRH encounters. Just over half of CRH users (403 of 804) had non-CRH cardiology encounters in addition to CRH encounters. In addition, there were 19,583 non-CRH users with 83,489 ambulatory encounters, meaning CRH users comprised 4% of the 20,387 total patients using ambulatory cardiology services in the region over the study period.

Among CRH users, 8% were women and 41% were ≥75 years old, compared to 5% and 49% respectively among non-CRH users. Similar proportions in both groups were rural or highly rural (26% for both CRH and non-CRH), highly-disabled (48% CRH, 47% non-CRH), and low-income (21% CRH, 20% non-CRH).

**Figure 1:**
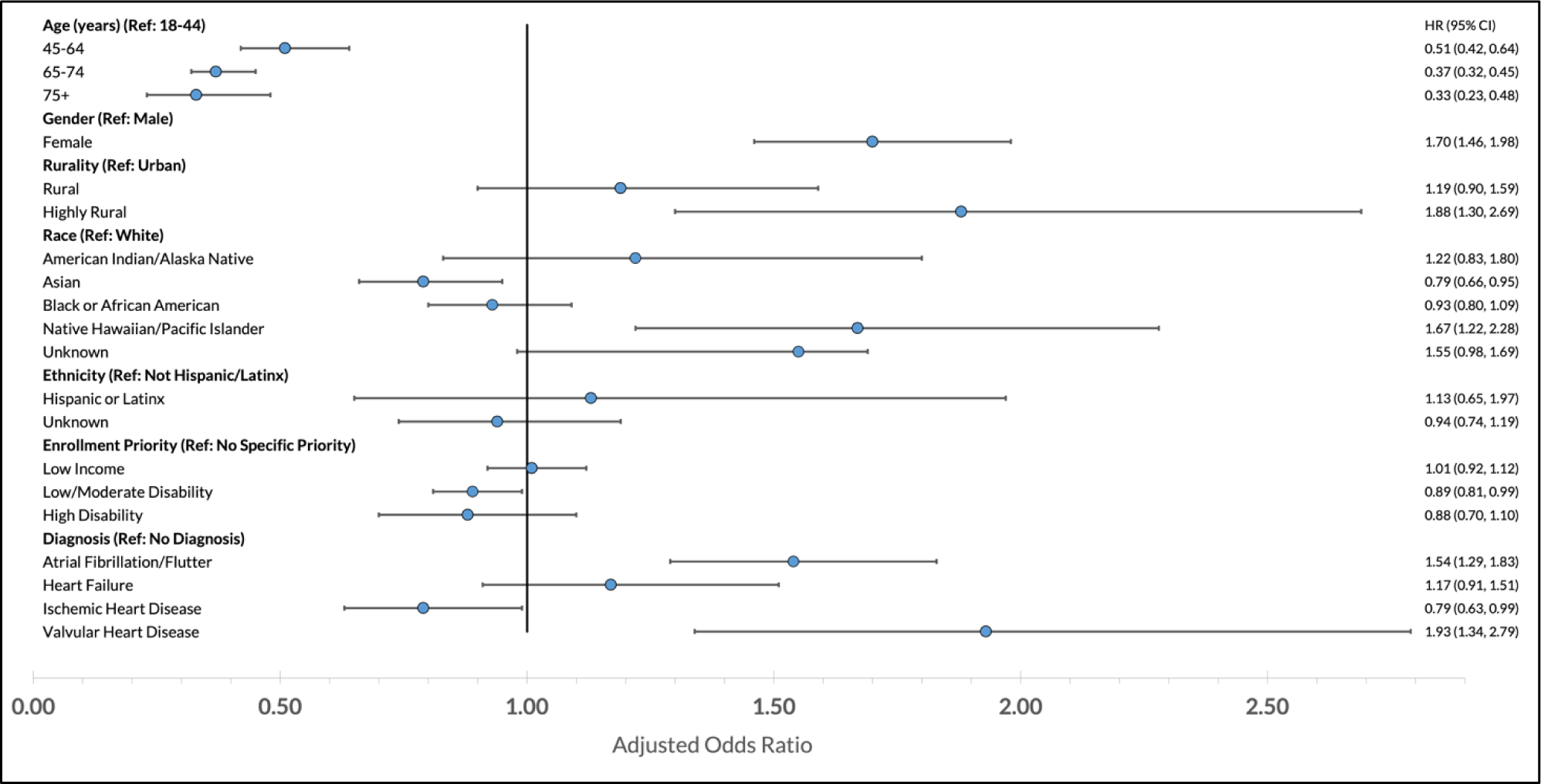
Adjusted Odds of Being a Clinical Resource Hub (CRH) Cardiology User Figure 1 represents the odds of using CRH adjusted for the sociodemographic and clinical characteristics shown and with clustering by the patient’s assigned primary care site.

In a multivariate logistic model with clustering at patient’s primary site, adjusted odds of using CRH were lower for older Veterans (AOR 0.33 for ≥75 [95% CI 0.23-0.48]) and higher for women (adjusted odds ratio [AOR] 1.70 [95% CI 1.46-1.98]). Highly rural Veterans also had higher adjusted odds of using CRH (AOR 1.88 [95% CI 1.30-2.69]). There were few significant differences by race/ethnicity, though patients of Native Hawaiian/Pacific Islander race had higher odds of using CRH (AOR 1.67 [95% CI 1.22-2.28]). Having a diagnosis of atrial fibrillation/flutter or valvular heart disease was also associated with higher adjusted odds of using CRH (AOR 1.54 [95% CI 1.29-1.83] and 1.93 [1.34-2.79], respectively).

The number of CRH patients increased over time (Figure 2), with some sites’ growth rates picking up more abruptly (e.g., Northern California) and others demonstrating a steadier increase (Sierra Nevada, Southern Nevada). Initial CRH encounters for these sites were 7/29/2021 (Southern Nevada), 9/8/2021 (Sierra Nevada), and 9/21/2021 (Northern California).

**Figure 2.**
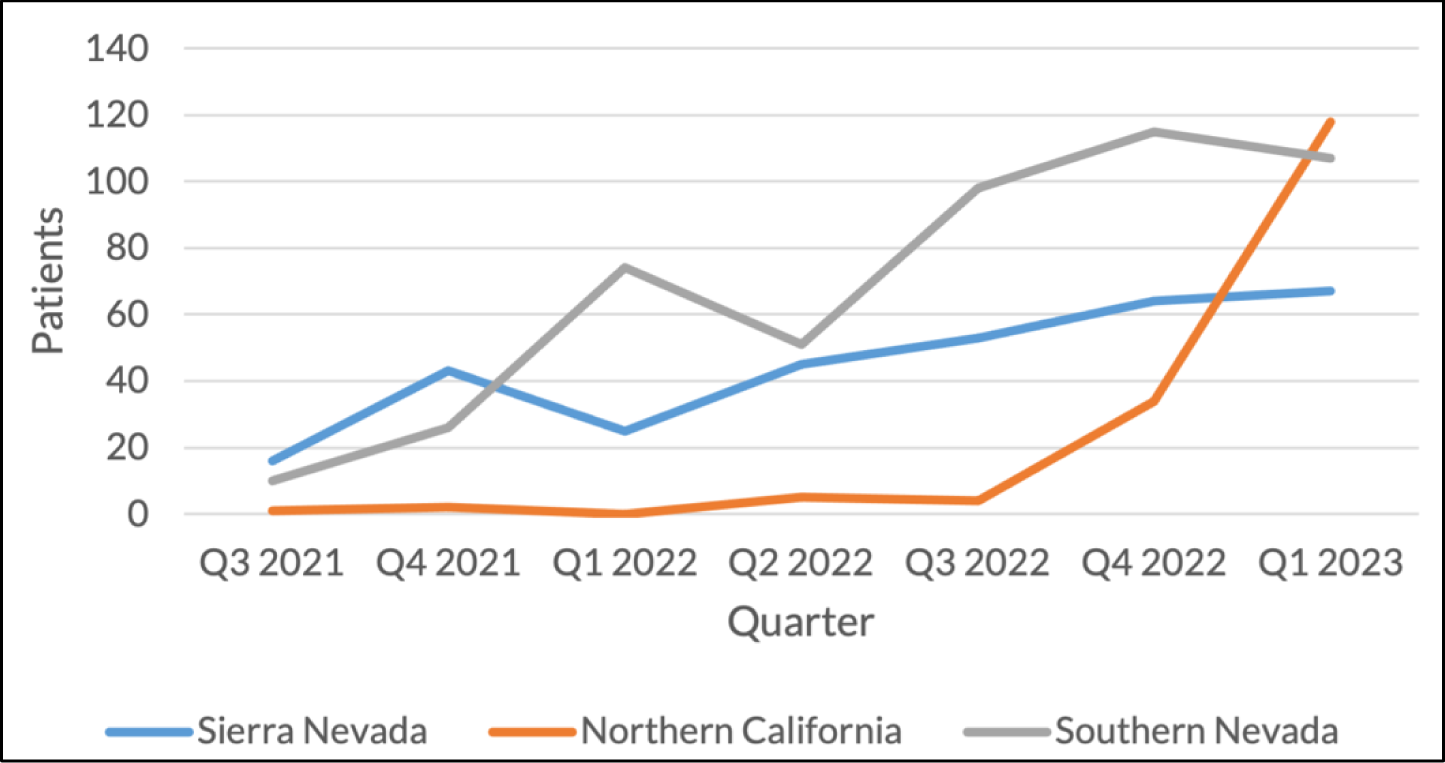
Unique CRH Patients at Implementing Sites Over Time (Total N=804; N across sites depicted=688) Figure 2 shows patients seen in CRH clinics each quarter by site over the study period.

### Encounters

The total number of ambulatory cardiology encounters in the region remained approximately constant over the study period (Figure 3); a slight uptick in total encounters took place in the first quarter of 2023, mostly due to an increase in in-person encounters. Figure S1 shows the breakdown in encounter modality among CRH sites over time. Figure 4 shows the growth in CRH cardiology encounters over time at the three sites with most CRH encounters (Sierra Nevada, or Reno; Southern Nevada, or Las Vegas; and Northern California, or Sacramento).

**Figure 3.**
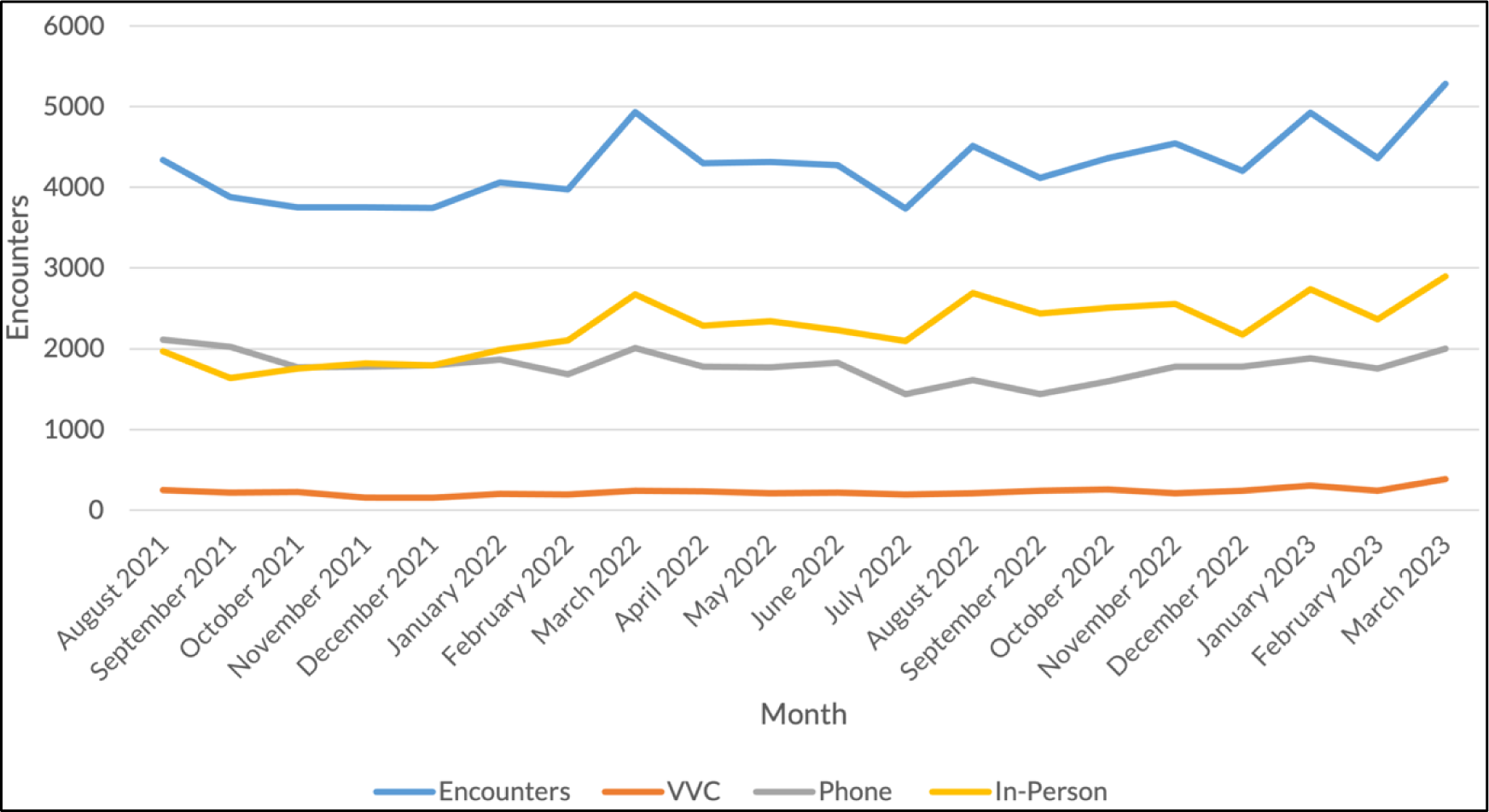
Cardiology Encounters Over Time by Encounter Modality, VHA Sierra Pacific Region^a^ Figure 3 depicts cardiology encounters in the VHA Sierra Pacific region (VISN 21) over the study period, including all CRH and non-CRH VA-based encounters.

**Figure 4.**
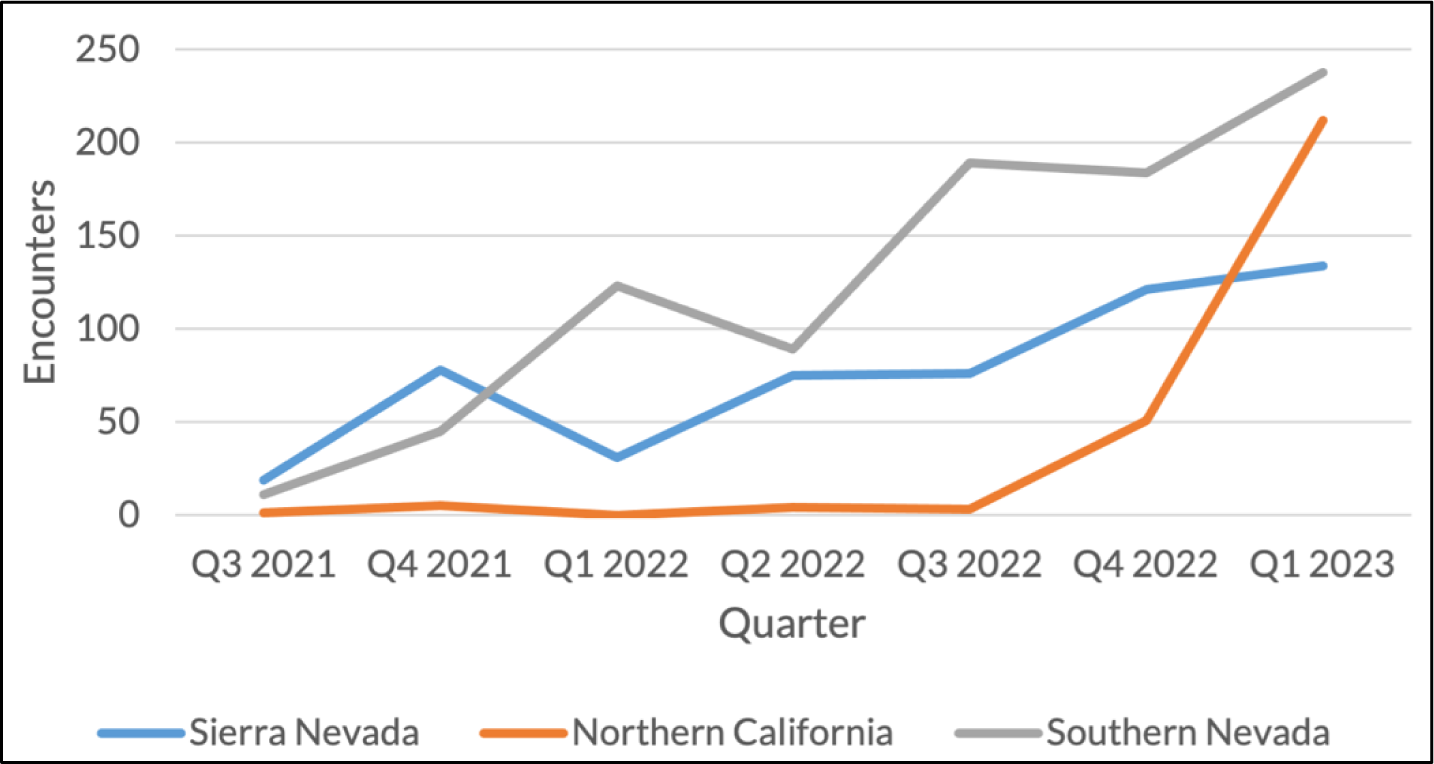
CRH Cardiology Encounters at Implementing Sites Over Time Figure 4 depicts CRH cardiology encounters in the VHA Sierra Pacific region (VISN 21) over the study period at implementing sites.

**Table 1.** Characteristics of Cardiology Patients, CRH Users (N=804) and Non-Users (N=19,583)

|  | CRH Users | CRH Non-Users | Effect Size Estimate<br>(Cramer's V or Cohen's d)<sup>a</sup> |
| --- | --- | --- | --- |
|  | N=804 | N=19,583 |  |
| **Used VVC<sup>b</sup>** | 438 (54.5) | 1,633 (8.3) | -0.81 |
| **Age, years (mean [SD])** | 69.5 (12.8) | 72.7 (11.1) | -0.05 |
| **Age, years, categorical** |  |  | 0.05 |
| 18-44 | 50 (6.2) | 515 (2.6) | - |
| 45-64 | 168 (20.9) | 2,998 (15.3) | - |
| 65-74 | 253 (31.5) | 6,470 (33.0) | - |
| 75+ | 333 (41.4) | 9,600 (49.0) | - |
| **Race** |  |  | 0.03 |
| American Indian or Alaska Native | 15 (1.9) | 275 (2.6) | - |
| Asian | 20 (2.5) | 765 (3.9) | - |
| Black or African American | 79 (9.8) | 2,011 (10.3) | - |
| Native Hawaiian or other Pacific Islander | 28 (3.5) | 503 (2.6) | - |
| Unknown/Missing | 87 (10.8) | 1,979 (10.1) | - |
| White | 574 (71.4) | 14,036 (71.7) | - |
| **Ethnicity** |  |  | 0.01 |
| Hispanic or Latino | 55 (6.8) | 1,569 (8.0) | - |
| Not Hispanic or Latino | 695 (86.4) | 16,837 (86.0) | - |
| Unknown/Missing | 54 (6.7) | 1,177 (6.0) | - |
| **Sex** |  |  |  |
| Female | 67 (8.3) | 840 (4.3) | -0.18 |
| Male | 737 (91.7) | 18,743 (95.7) | 0.01 |
| **Rurality** |  |  | 0.02 |
| Urban | 592 (73.6) | 14,574 (74.4) | - |
| Rural | 197 (24.5) | 4,745 (24.2) | - |
| Highly Rural | 8 (1.0) | 191 (1.0) | - |
| Missing | 7 (0.9) | 74 (0.4) | - |
| **Enrollment Priority** |  |  | 0.02 |
| No special priority | 113 (14.1) | 2,650 (13.5) | - |
| Low/moderate disability | 137 (17.0) | 3,644 (18.6) | - |
| High disability | 387 (48.1) | 9,246 (47.2) | - |
| Low income | 165 (20.5) | 3,941 (20.1) | - |
| Missing | 2 (0.2) | 102 (0.5) | - |
| **Diagnoses** |  |  |  |
| Atrial Fibrillation/Flutter | 320 (39.8) | 6,300 (32.2) | -0.02 |
| Heart Failure | 153 (19.0) | 3,405 (17.4) | -0.10 |
| Ischemic Heart Disease | 261 (32.5) | 8,137 (41.6) | -0.17 |
| Valvular Heart Disease | 161 (20.0) | 2,530 (12.9) | 0.12 |

<sup>a</sup> Standardized mean differences calculated via Cramer's V for categorical variables and via Cohen's d for continuous variables and binary categorical variables.
<sup>b</sup> Indicates use of VVC during study period.

#### Modality

714 of the 1,961 CRH encounters (36%) were conducted via video, with the remainder (1,247, or 64%) conducted via telephone. For non-CRH encounters, 4% were conducted via video (N=3,830), 42% via telephone (N=34,757), and just over half, or 54%, occurred in person (N=44,902).

Ever using CRH was associated with much higher adjusted odds of ever using video care (AOR 29.73 [95% CI 11.38-77.66]) (Table 2). Age was associated with lower adjusted odds of video care use according to a gradient, with an AOR of 0.44 (95% CI 0.37-0.53) for Veterans 75 years or older. Living in a rural location was associated with higher adjusted odds of video care use (AOR 1.27 [95% CI 1.06-1.52]), though this finding was not significant for those in highly rural locations (AOR 1.30 [95% CI 0.82-2.05]).

**Table 2.** Adjusted Odds of Using Video Care During Study Period Among All Cohort Patients (N=20,387)

|  | Adjusted Odds Ratio | 95% Confidence Interval |
| --- | --- | --- |
| **CRH User Status** |  |  |
| Never used CRH | (ref) | (ref) |
| Used CRH | 29.73 | (11.38, 77.66) |
| **Age, years, categorical** |  |  |
| 18-44 | (ref) | (ref) |
| 45-64 | 0.80 | (0.72, 0.89) |
| 65-74 | 0.57 | (0.49, 0.66) |
| 75+ | 0.44 | (0.37, 0.53) |
| **Race** |  |  |
| American Indian or Alaska Native | 1.27 | (0.96, 1.67) |
| Asian | 0.68 | (0.50, 0.92) |
| Black or African American | 0.78 | (0.48, 1.25) |
| Native Hawaiian or other Pacific Islander | 0.83 | (0.64, 1.08) |
| Unknown | 0.90 | (0.79, 1.03) |
| White | (ref) | (ref) |
| **Ethnicity** |  |  |
| Hispanic or Latino | 0.91 | (0.68, 1.22) |
| Not Hispanic or Latino | (ref) | (ref) |
| Unknown | 1.12 | (0.92, 1.37) |
| **Gender** |  |  |
| Female | 1.20 | (1.01, 1.42) |
| Male | (ref) | (ref) |
| **Rurality** |  |  |
| Urban | (ref) | (ref) |
| Rural | 1.27 | (1.06, 1.52) |
| Highly Rural | 1.30 | (0.82, 2.05) |
| Missing | 1.81 | (0.65, 5.04) |
| **Enrollment Priority** |  |  |
| No special priority | (ref) | (ref) |
| Low/moderate disability | 1.02 | (0.87, 1.21) |
| High disability | 1.13 | (0.93, 1.36) |
| Low income | 0.81 | (0.70, 0.94) |
| Missing | 0.54 | (0.25, 1.13) |
| **Diagnoses** |  |  |
| Atrial Fibrillation/Flutter | 1.23 | (1.09, 1.40) |
| Heart Failure | 1.37 | (0.92, 2.03) |
| Ischemic Heart Disease | 1.25 | (1.01, 1.54) |
| Valvular Heart Disease | 1.70 | (1.30, 2.21) |

## Discussion

In just under two years of operation, this hub-and-spoke, primarily virtual cardiology clinic in VHA’s Sierra Pacific region served over 800 Veterans hailing from across the region in nearly 2,000 virtual encounters for evaluation and management of cardiovascular disease. The CRH program served women and highly-rural-dwelling Veterans at higher rates and similar proportions of highly-disabled and low-income Veterans compared to conventional cardiology clinics in the same region; conversely, the CRH patient population skewed younger than the conventional VHA clinic population. This suggests that such a predominately-virtual model of specialty care may be an effective method for accessing care for many high-need groups, although more targeted efforts may be required to reach older Veterans.

The reporting of these early results coincides with a shift in virtual care use from effectively a requirement during the national emergency phase of the COVID-19 pandemic, to an option–for patients and clinicians alike.^13^ This evolution brings both an opportunity and a mandate for rigorous study of how, when, and for whom virtual care should be employed, and how telehealth visits affect quality of care, resource use, and health outcomes.^14^ The current study is formative, with a focus on examining patterns of use; this lays a foundation for follow-up studies delving into the latter set of questions.

The concern of the digital divide^15^ is ever-present when considering use of virtual care: will a primarily virtual model of care inadvertently exclude groups frequently falling on the wrong side of the divide, such as those who are rural-dwelling or low-income? Based on these findings, this particular program has reached historically marginalized groups in VHA, such as women, racial/ethnic minority Veterans, or those who are highly disabled or low-income, at similar or higher rates than the conventional model. A notable exception is among older individuals, who used CRH at much lower rates than their younger counterparts. The majority of older individuals in the United States are interested in conducting visits via telehealth,^16^ yet disparities in use by age have been widely demonstrated in VHA both in general and in cardiology.^6,7^ Establishing the source(s) of this discrepancy—whether due to true differences in interest in receiving care via a primarily-virtual care model, lower rates of offering the CRH program to older individuals, familiarity with navigating virtual technologies, or other factors—will be an important focus of follow-up work.

We found that patients with diagnoses of atrial fibrillation/flutter or valvular heart disease had higher adjusted odds of being CRH users, unlike patients with diagnoses of heart failure or ischemic heart disease. This finding may reflect program-specific offerings (for example, clinics or physicians in the hub site with particular expertise in managing these conditions, or alternatively, a perceived lack of capacity to manage them at spoke sites), or a sense that these conditions are more amenable to primarily virtual management. Planned qualitative work, including interviews with program clinicians and administrators, will help to differentiate between these possible drivers.

For virtual care models designed to improve patient access to a given service, it is essential to establish whether that model offloads the conventional model, as intended, or simply induces more demand (e.g., patients whose cardiovascular diseases would have otherwise been cared for in a primary care setting are instead referred for cardiology care). While the current study does not aim to definitively answer this question, the fact that total cardiology encounters remained constant in the region over the study period suggests that there was not a strong demand-creation effect of the CRH model. However, to date CRH patients comprise only a small fraction of total regional patients using cardiology services, so continued attention to this question will be important as the program grows.

### Limitations

Within the current data and study design, we are limited in interpretation of various aspects of our findings. For example, although we can capture which CRH users have also used conventional care, our data lacks the granularity to understand how and when this is the case; subsequent qualitative work will further elucidate these care patterns. At present our data is limited to encounters within VHA and does not extend to VA-purchased care in the community, meaning we cannot fully conclude whether CRH affects consumption of this costly form of care. This question, and characterization of other important facets of care associated with the program, such as patient, caregiver, and clinician satisfaction, clinical outcomes, and drivers of more and less successful program implementation, are left for future work. Finally, this analysis focused on a particular region and healthcare system, and therefore may not be fully generalizable to other healthcare settings—although as Burnett et al (2023) note in their publication on the early CRH implementation experience, “…some CRH design elements and experiences are unique to the VHA system, [but] overall experience with telehealth hubs—including attempts to improve capacity for service provision, increase access, and deployment of telehealth services—is likely highly relevant to other health care systems.”^8^

## Conclusions

The Cardiology CRH program represents a virtual-predominant model of care, implemented over a relatively short period of time, that has served over 800 patients to date in the VHA region serving Northern California, Nevada, and the Pacific Islands. These data from the first two years of program implementation suggest that the program reached many of the most historically marginalized sub-populations of Veterans, including women, rural-dwellers, and low-income Veterans, at similar or higher rates compared to conventional cardiology care in the region. A notable exception was older individuals, who used CRH care at much lower rates; further work will examine the extent to which patient preference versus other factors drove this dynamic.

## Data Availability

Data is not publicly available per VA policy, but relevant code will be shared with VA-based researchers upon request.

## Acknowledgments

The authors thank the Virtual Access QUERI team for supporting this evaluation, including Cindie Slightam, MPH and Camila Chaudhary, MPH, from VA Palo Alto Health Care System, for providing project management support and Liberty Greene, MEd and James Van Campen, MS for data consulting. Preliminary results of this work will be presented at the American Heart Association Scientific Sessions, November 13, 2023.

